# Rhinovirus in adults with severe lower respiratory tract infection: incidental carriage or primary pathogen? A comparative study across different clinical status categories

**DOI:** 10.1101/2025.11.14.25340248

**Authors:** Ana Helena Perosa, Klinger S. Faico-Filho, Karen Pendeloski, Rodrigo Matheus, Nancy Bellei

## Abstract

**Background:** Rhinovirus (RV) is a recognized respiratory pathogen, but its role in severe lower respiratory tract infections (LRTI) in adults remain debated. The aim of the study was to investigate RV in different clinical status categories in a tertiary hospital.

**Methods:** We conducted a cross-sectional study between January 2023 and August 2025, enrolling 2,688 participants: asymptomatic adults (n=193), adults outpatients with acute respiratory infection (ARI) (n=1,274), hospitalized adults with severe LRTI (n=809), and hospitalized children with LRTI (n=412). RV detection was performed by RT-qPCR.

**Results:** RV detection rates were 4.7% in asymptomatic adults, 7.8% in hospitalized adults, 15.6% in adults outpatients and 20.9% in hospitalized children. The detection rate in hospitalized adults was statistically similar to that of asymptomatic adults (p=0.088), but significantly lower than that of ambulatory adults (p<0.001) and hospitalized children (p<0.001). Among RV-positive hospitalized adults, 90.5% had underlying comorbidities, with 65.1% presenting immunosuppressive conditions.

**Conclusions:** In hospitalized adults with LRTI, RV detection rates are comparable to those in asymptomatic individuals, suggesting frequent incidental carriage rather than a primary causal link to severe disease. Conversely, RV is a major pathogen in acute, non-severe community illness in adults and a clearly pathogenic agent in severe LRTI in children.

## Introduction

Rhinovirus (RV) has long been recognized as a ubiquitous respiratory pathogen, traditionally associated with mild upper respiratory tract infections but increasingly implicated in severe lower respiratory tract disease (LRTI), particularly in vulnerable populations ^(1,2)^. While RV is a well-established cause of severe disease in children, its role in severe LRTI requiring hospitalization in adults is less clear. The conventional paradigm often attributes severe illness to any detected respiratory pathogen, but this may lead to misdiagnosis, delayed treatment for alternative causes, and inappropriate use of antibiotics. Previous studies suggested that the presence of RV in hospitalized adults may not be a strong indicator of a causal link to severe disease, especially when other pathogens are also detected ^(3,4)^.

To address this ambiguity, we conducted a study in Sao Paulo, Brazil, with the primary objective of comparing RV detection rates across different clinical status categories of patient’s presentations. By analyzing samples from asymptomatic individuals, adults outpatients with acute illness, hospitalized adults with severe LRTI, and hospitalized children, we aimed to provide a comprehensive perspective on the pathogenic role of RV in these diverse populations. Our hypothesis was that RV detection in hospitalized adults would more closely resemble that of asymptomatic individuals than that of hospitalized children, supporting the notion of incidental carriage.

## Materials and Methods

This was a cross-sectional study conducted in Sao Paulo, Brazil, between January 2023 and August 2025. A total of 2,688 participants were enrolled across four distinct clinical groups:

- Hospitalized adults: 809 patients (aged >12 years) admitted with LRTI.
- Hospitalized children: 412 patients (aged ≤12 years) admitted with LRTI.
- Ambulatory adults: 1,274 healthcare workers presenting with symptoms of acute respiratory infection (ARI) at an outpatient clinic.
- Asymptomatic adults: 193 individuals undergoing routine pre-surgical screening with no signs or symptoms of respiratory illness.

Nasopharyngeal swab samples were collected from all participants and placed in 2mL of sterile Ringer’s lactate. Total nucleic acid was extracted using Extracta Kit Fast - DNA e RNA Viral (Loccus, Brazil), according to the manufacturer’s instructions. RV detection was performed by a one-step Real-time RT-qPCR with oligonucleotides previously described ^(5)^, using AgPath-ID™ One-Step RT-PCR Reagents (Applied Biosystems, USA) with 5µL of purified RNA, 800nM of each primer and 200nM of the TaqMan probe. The reaction was performed on a Quantstudio 6 Pro Real-Time PCR System (Applied Biosystems) for 10 min at 50°C and 10 min at 95°C, followed by 45 cycles of 15s at 95°C, and 30s at 55°C (data collection). Samples with Ct ≤40 were considered positive.

Descriptive statistics were used to summarize the demographic and clinical characteristics of the study participants. The chi-square test was used to compare the RV detection rates between groups and a □-value of <0.05 was considered statistically significant. All statistical analyses were performed using SPSS version 26.

This study protocol was approved by the Research Ethics Committee of Sao Paulo Hospital and the Federal University of Sao Paulo (CEP/UNIFESP n° 5505, CAAE n° 74157223.2.0000.5505).

## Results

A total of 2,688 participants were included in the analysis: 2,276 adults (median 48 years, IQR 35-59) and 412 children (median 2 years, IQR 0.6-6). The overall RV detection rate was 13.3% (357/2,688), with 15.6% in adults outpatients, 7.8% in hospitalized adults, 4.7% in asymptomatic adults, and 20.9% in hospitalized children. The clinical characteristics and RV positivity of each group are detailed in table I.

**Table I.** Rhinovirus detection rates across clinical groups.

| **Clinical Group** | **Total (n)** | **RV Positive** | **Clinical Significance** |
| --- | --- | --- | --- |
| Asymptomatic adults | 193 | 9 (4.7%) | Baseline carriage |
| Hospitalized adults with LRTI | 809 | 63 (7.8%) | Similar to asymptomatic* |
| Adults outpatients with ARI | 1,274 | 199 (15.6%) | Likely pathogenic |
| Hospitalized children with LRTI | 412 | 86 (20.9%) | Clearly pathogenic |

LRTI: Lower Respiratory Tract Infection; ARI: Acute Respiratory Infection; RV: rhinovirus. \* $p=0.088$

### Rhinovirus in Adults

The most remarkable finding of our study lies in the similarity between RV detection rates in asymptomatic individuals (4.7%) and hospitalized adults with severe LRTI (7.8%) (□=0.088). The detection rate in ambulatory adults with ARI was significantly higher than in hospitalized adults (□<0.001) and asymptomatic adults (□<0.001). The data shows that ambulatory adults were almost twice as likely to test positive for RV. The highest RV positivity was found in ambulatory patients, with a rate of 24.1% in the 70-79 years age group. This is higher than the rate for hospitalized patients in the same age bracket, which was 5.2% (table II).

**Table II.** Rhinovirus detection according to age and clinical presentation in adults.

| **Age** | **Total** | **Hospitalized** | **Outpatients** | **Asymptomatic** |
| --- | --- | --- | --- | --- |
| **Age (years)** | 13-99 | 13-99 | 18-81 | 13-94 |
| **Average (SD)** | 47.3 (16.5) | 50.4 (19.9) | 45.4 (13.4) | 46.9 (17.8) |
| **Median** | 48 | 54 | 47 | 46 |
| **13-49 years** | 178/1,187 (15%) | 31/350 (8.9%) | 139/734 (18.9%) | 8/103 (7.8%) |
| **50-59 years** | 55/521 (10.5%) | 12/133 (9%) | 43/352 (12.2%) | 0/36 |
| **60-69 years** | 21/374 (5.6%) | 11/184 (6%) | 10/158 (6.3%) | 0/32 |
| **70-79 years** | 13/162 (8%) | 6/116 (5.2%) | 7/29 (24.1%) | 0/17 |
| **≥ 80 years** | 4/32 (12.5%) | 3/26 (11.5%) | 0/1 | 1/5 (20%) |
| **Total** | 2,276 | 809 | 1,274 | 193 |

Further analysis of the 63 hospitalized adults with positive RV tests revealed a high prevalence of comorbidities (90.5%, 57/63) with immunosuppressive conditions as the most common (65.1%, 41/63). Cardiovascular conditions were reported in 19% (12/63), respiratory issues in 11.1% (7/63), and other conditions like rheumatological, renal, hepatic and metabolic were found in 36.5% (23/63) of patients. Severe outcomes were observed, with 42.8% (27/63) of these patients requiring an ICU stay, 20.6% (13/63) needing tracheal intubation, and 17.5% (11/63) resulting in death.

### Rhinovirus in Children

RV detection in hospitalized children was 20.9% (86/412), which is higher compared to all adult groups (□<0.001). There is no statistically significant difference in the RV detection rates across the different age groups (table III).

**Table III.** Characteristics and RV detection in hospitalized children by age groups.

| Age | Total (n=412) | RV Positive |
| --- | --- | --- |
| **Mean (SD)** | 3.4 (3.6) | 3.7 (3.7) |
| **Median** | 2 | 2 |
| **Age groups** |  |  |
| 0-6 months | 95 | 19 (20%) |
| 7-11 months | 47 | 6 (12.8%) |
| 1-2 years | 91 | 20 (22%) |
| 3-5 years | 75 | 17 (22.7%) |
| 6-12 years | 104 | 24 (23.1%) |

Among children with RV infection, 61.6% (53/86) had comorbidities, with respiratory issues reported in 26.7% (23/86), cardiovascular conditions in 23.2% (20/86), immunosuppressive conditions in 5.8% (5/86) and other diseases like rheumatological, renal, hepatic, and metabolic reported in 30.2% (26/86). Severe outcomes were also observed, with 22.1% (19/86) of these children admitted to the ICU and 7% (6/86) requiring mechanical ventilation.

RV was detected throughout all months during the study period, with no seasonal pattern. While there were peaks in positivity, such as 23% in June 2023, 23.4% in January 2024 and 27.6% in August 2025, no consistent pattern was observed over study period (figure 1).

**Figure 1:**
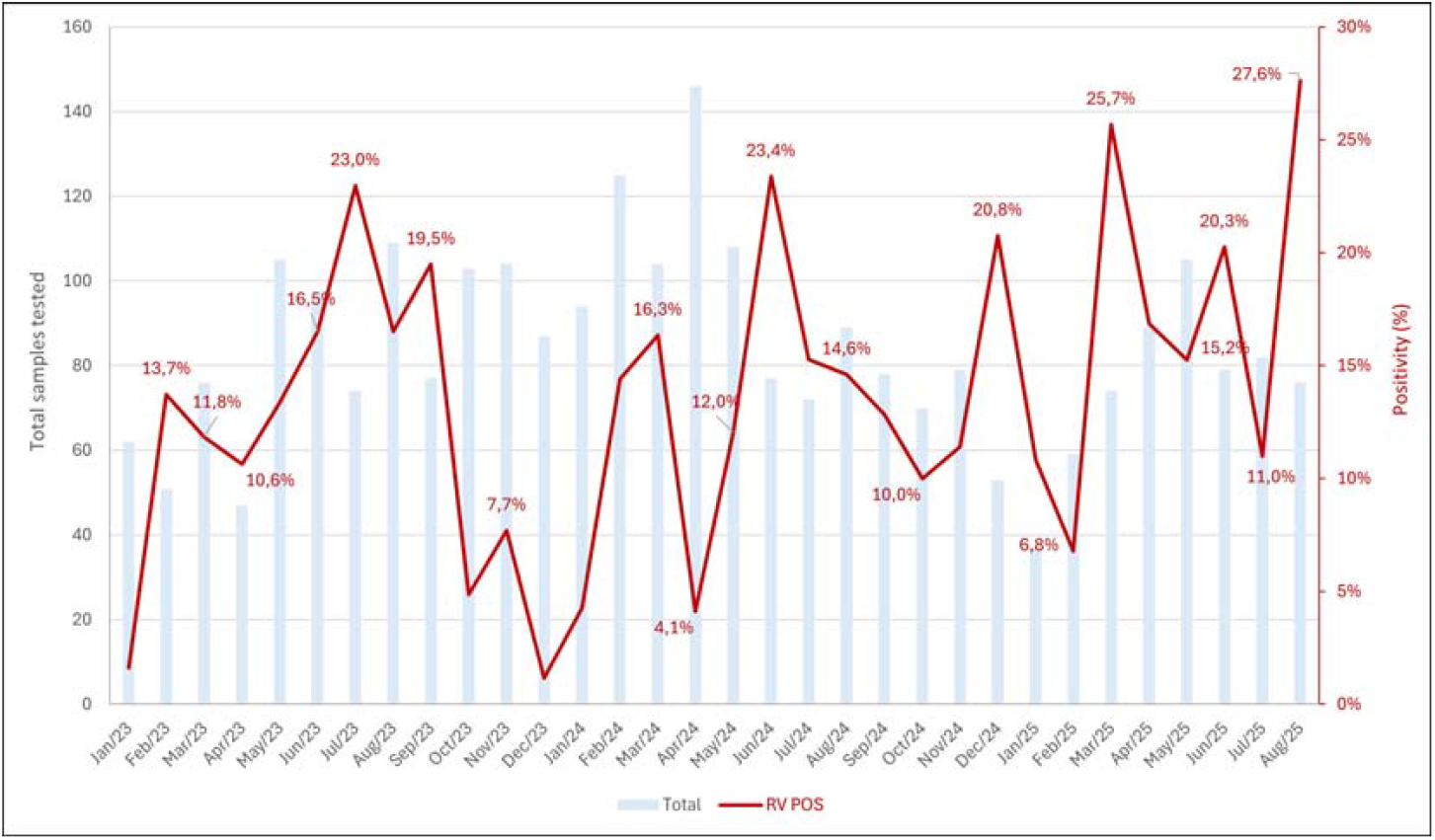
Monthly distribution of collected samples and RV positivity rate from January 2023 to August 2025. The blue bars represent the number of patients included, while the red line indicates the percentage of RV positive cases.

## Discussion

In this cross-sectional study, we investigated the pathogenic role of RV by comparing detection rates across four distinct groups: asymptomatic adults, adults outpatients with ARI, hospitalized adults with LRTI, and hospitalized children with LRTI. The most notable finding was the similar RV detection rate between asymptomatic adults (4.7%), and hospitalized adults with severe LRTI (7.8%) (□=0.088). This finding strongly suggests that, for many hospitalized adults, RV presence may be a coincidental finding rather than the primary driver of their severe disease. Some biological and clinical factors support this hypothesis. For example, adult immune systems - having encountered multiple RV strains throughout life - may sustain prolonged viral shedding patterns without active disease manifestation ^(6,7)^.

Additionally, an analysis of severely ill RV-positive adults revealed a very high prevalence of comorbidities (90.5%), particularly immunosuppressive conditions (65.1%). These findings support the clinical recommendation to exercise caution in attributing severe adult LRTI solely to RV and to aggressively investigate other potential causes, especially in the context of high-risk comorbidities. While RV may contribute to illness severity, pre-existing conditions likely played a crucial role in determining ICU admission (42.8%) and fatal outcomes (17.5%). This underscores the multifactorial nature of disease in adults, with individual host factors being especially critical. Additionally, the attribution of RV as a direct cause of LRTI based solely on upper respiratory tract samples remains controversial. Despite the high concordance between upper and lower respiratory tract specimens ^(8)^, RV detection in the upper respiratory tract alone is insufficient to confirm etiologic significance in adults.

In contrast, the RV detection rate in adults outpatients (15.6%) was significantly higher than in both asymptomatic and hospitalized adults (p<0.001), suggesting that RV is indeed a major pathogen of acute, non-severe respiratory illness in the adult community. Interestingly, the highest RV positivity in adults outpatients was observed in the 70–79 age group (24.1%), significantly exceeding the rate in their hospitalized counterparts (5.2%). This further underscores the difference in RV’s role: causing mild-to-moderate illness in the community versus being a less frequent primary cause of severe illness requiring hospitalization even in the elderly.

The data for hospitalized children were different. Their RV detection rate was significantly higher than that of all adult groups (□<0.001), aligning with the established view that RV is a clear major pathogen responsible for severe LRTI in children. For children, though comorbidities were also present (61.6%), the substantially higher detection rate strongly suggests a more direct pathogenic link.

Finally, the continuous detection of RV throughout the study period, without a clear seasonal pattern, suggests it is an endemic, year-round circulating pathogen in Sao Paulo, Brazil, similar to observations in other tropical and subtropical regions ^(9,10)^.

A limitation of this study was the lack of RV species identification through sequencing, which could have impacted our results, as some studies suggest that RV-B is associated with lower pathogenicity ^(11,12)^. On the other hand, other studies have not demonstrated a consistent association between specific RV species and disease severity ^(3)^.

This study provides epidemiological evidence distinguishing the roles of RV across various patient populations in São Paulo, Brazil. In hospitalized adults with severe LRTI, RV detection rates are comparable to those in asymptomatic adults, suggesting that RV is frequently an incidental carriage, not the sole cause of severe disease. The severity and poor outcomes observed in RV-positive hospitalized adults are driven by an overwhelming burden of comorbidities, particularly immunosuppression. RV acts as a major pathogen in acute, non-severe community illness in adults and is clearly a pathogenic agent in severe LRTI in hospitalized children. These findings call for a fundamental reassessment of how we interpret RV detection in hospitalized adults and emphasize the need for continued diagnostic vigilance to identify alternative pathogens that may require specific therapeutic interventions. As we advance toward more personalized and precise medical care, the nuanced interpretation of respiratory pathogen detection becomes increasingly critical for optimizing patient outcomes and ensuring appropriate antimicrobial stewardship in the hospital setting.

## Data Availability

All data produced in the present study are available upon reasonable request to the authors.

